# Tryptophan metabolism determines outcome in tuberculous meningitis: a targeted metabolomic analysis

**DOI:** 10.1101/2023.01.08.23284316

**Authors:** Edwin Ardiansyah, Julian Avila Pacheco, Le Thanh Hoang Nhat, Sofiati Dian, Dao Nguyen Vinh, Hoang Thanh Hai, Kevin Bullock, Bachti Alisjahbana, Mihai G Netea, Riwanti Estiasari, Trinh Thi Bich Tram, Joseph Donovan, Dorothee Heemskerk, Tran Thi Hong Chau, Nguyen Duc Bang, Ahmad Rizal Ganiem, Rovina Ruslami, Valerie ACM Koeken, Raph L Hamers, Darma Imran, Kartika Maharani, Vinod Kumar, Clary B. Clish, Reinout van Crevel, Guy Thwaites, Arjan van Laarhoven, Nguyen Thuy Thuong Thuong

**Affiliations:** Research Center for Care and Control of Infectious Diseases, Universitas Padjadjaran, Bandung, Indonesia; Department of Internal Medicine and Radboud Center of Infectious Diseases (RCI), Radboud University Medical Center, Nijmegen, Netherlands; The Broad Institute of MIT and Harvard, Cambridge, MA, USA; Oxford University Clinical Research Unit, Ho Chi Minh City, Vietnam; Department of Internal Medicine, Hasan Sadikin Hospital, Faculty of Medicine, Universitas Padjadjaran, Bandung, Indonesia; Department of Neurology, Hasan Sadikin Hospital, Faculty of Medicine, Universitas Padjadjaran, Bandung, Indonesia; Department of Neurology, Cipto Mangunkusumo Hospital, Faculty of Medicine Universitas Indonesia; Centre for Tropical Medicine and Global Health, Nuffield Department of Medicine, University of Oxford, Oxford, United Kingdom; London School of Hygiene and Tropical Medicine, Keppel St, London, United Kingdom; Department of Medical Microbiology and Infection Prevention, Amsterdam University Medical Centre, Amsterdam, the Netherlands; Hospital for Tropical Diseases, District 5, Ho Chi Minh City, Vietnam; Pham Ngoc Thach Hospital for Tuberculosis and Lung Disease, District 5, Ho Chi Minh City, Vietnam; Department of Biomedical Science, Faculty of Medicine, Universitas Padjadjaran, Bandung, Indonesia; Department of Computational Biology for Individualised Infection Medicine, Centre for Individualised Infection Medicine (CiiM) & TWINCORE, joint ventures between the Helmholtz-Centre for Infection Research (HZI) and the Hannover Medical School (MHH), 30625 Hannover, Germany; Oxford University Clinical Research Unit Indonesia, Faculty of Medicine Universitas Indonesia

## Abstract

**Background:** Cellular metabolism is critical for the host immune function against pathogens, and metabolomic analysis may help understand the characteristic immunopathology of tuberculosis. We performed targeted metabolomic analyses in a large cohort of patients with tuberculous meningitis (TBM), the most severe manifestation of tuberculosis, focusing on tryptophan metabolism.

**Methods:** We studied 1069 Indonesian and Vietnamese adults with TBM (26.6% HIV-positive), 54 non-infectious controls, 50 with bacterial meningitis, and 60 with cryptococcal meningitis. Tryptophan and downstream metabolites were measured in cerebrospinal fluid (CSF) and plasma using targeted liquid chromatography mass-spectrometry. Individual metabolite levels were associated with survival, clinical parameters, CSF bacterial load and 92 CSF inflammatory proteins.

**Results:** CSF tryptophan was associated with 60-day mortality from tuberculous meningitis (HR=1.16, 95%CI=1.10-1.24, for each doubling in CSF tryptophan) both in HIV-negative and HIV-positive patients. CSF tryptophan concentrations did not correlate with CSF bacterial load nor CSF inflammation but were negatively correlated with CSF interferon-gamma concentrations. Unlike tryptophan, CSF concentrations of an intercorrelating cluster of downstream kynurenine metabolites did not predict mortality. These CSF kynurenine metabolites did however correlate with CSF inflammation and markers of blood-CSF leakage, and plasma kynurenine predicted death (HR 1.54, 95%CI=1.22-1.93). These findings were mostly specific for TBM, although high CSF tryptophan was also associated with mortality from cryptococcal meningitis.

**Conclusion:** TBM patients with a high baseline CSF tryptophan or high systemic (plasma) kynurenine are at increased risk of mortality. These findings may reveal new targets for host-directed therapy.

**Funding:** This study was supported by National Institutes of Health (R01AI145781) and the Wellcome Trust (110179/Z/15/Z and 206724/Z/17/Z).

## Introduction

Tuberculous meningitis (TBM) is the most severe manifestation of tuberculosis affecting approximately 160,000 adults each year.^1^ Patients suffer from varying degrees of intracerebral inflammation, commonly manifest as leptomeningitis, vasculitis and space-occupying brain lesions (tuberculomas). Hydrocephalus, stroke, seizures, focal neurological deficits, and loss of consciousness are common complications and lead to death in around 30% of patients, even when treated with anti-tuberculosis drugs and adjuvant corticosteroid therapy.^1^ Development of more effective host-directed therapy is hampered by a lack of knowledge on the biological pathways involved in the immunopathology of TBM.^2^

Metabolism is critical for the function of immune cells, and analysis of cerebrospinal fluid (CSF) metabolites could help unravel underlying biological mechanisms in TBM. Previously, using a large-scale metabolomics analysis, we found that lower cerebrospinal fluid (CSF) tryptophan concentrations were associated with survival of TBM patients in Indonesia.^3^ This study did not include HIV-infected patients and the association was not validated in other populations.^4,5^ Moreover, there is a need to investigate the downstream metabolites in the kynurenine pathway (**Figure 1**), through which 95% of tryptophan is initially catabolized via indoleamine 2,3-dioxygenase (IDO) or tryptophan 2,3-dioxygenase (TDO) and which includes metabolites with putative neuroprotective (e.g. kynurenic acid) or neurodamaging (e.g. quinolinic acid) properties.^6^ Lastly, there is a need to compare these findings in other neuro-infectious diseases to distinguish disease specific from broader mechanisms.

**Figure 1.**
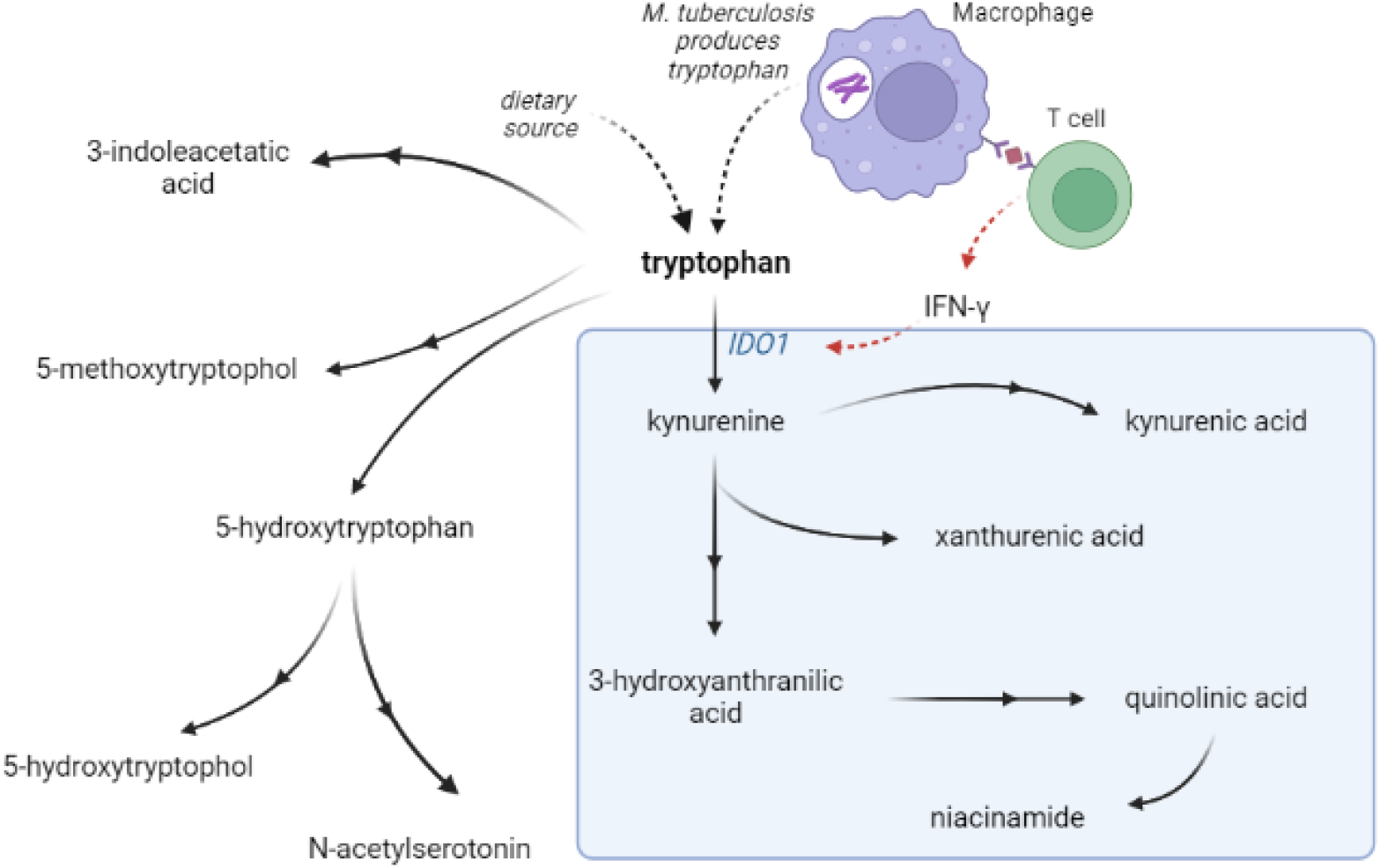
Tryptophan metabolism pathway. Tryptophan is metabolized mainly through the kynurenine pathway through indoleamine 2,3-dioxygenase 1 (IDO1), generating kynurenine and its downstream metabolites (blue box). IDO1 is partly stimulated by M. tuberculosis-induced interferon gamma (IFN-y) production by T helper 1 cells.

We therefore sought to define and validate the relationship between tryptophan and its metabolites and survival from TBM in large, independent populations, including HIV-positive individuals. We aimed to confirm that a higher CSF tryptophan would predict higher mortality across different populations and we hypothesized that high tryptophan would be associated with a higher CSF bacterial load, more inflammation and lower downstream kynurenine metabolites. We lastly sought to investigate how systemic (plasma) metabolite concentrations linked to outcome.

## Materials and Methods

### Setting and Patients

Patients with subacute meningitis were included from the Hospital for Tropical Diseases and Pham Ngoc Thach Hospital for Tuberculosis and Lung Disease in Ho Chi Minh City, Vietnam between 2011-2014,^4,7^ and Hasan Sadikin hospital in Indonesia between 2007-2019.^3,5^ TBM patients were defined as having ‘definite TBM’ if they had either microbial confirmation by Ziehl-Neelsen staining, positive CSF culture or GeneXpert. Based on previous studies,^5^ probable TBM was defined as clinically suspected TBM fulfilling at least 2 out of the following 3 criteria: CSF leukocytes ≥ 5 cells/μL, CSF/blood glucose ratio < 0.5, and CSF protein > 0.45 g/L. Patients were treated with antibiotics according to national guidelines for 180 days minimally, and received adjunctive dexamethasone starting at 0·3 mg/kg for grade I and 0·4 mg/kg for grade II or III tuberculous meningitis and tapered thereafter.^8^ Patients were followed-up clinically or by phone up until day 180 from admission. Primary outcome was 60-day survival, when most deaths attributable to TBM occur. As a secondary endpoint, earlier and later mortality were explored separately. We ensured equal power for both time windows by separating them by the median time to death for those patients who died during the total follow up of 180 days.

Patients without an infection (non-infectious controls) were included from the same sites. In Indonesia, patients in this group had undergone a lumbar puncture for suspected central nervous system infection or subarachnoid bleeding, but infection was excluded by negative microscopy, GeneXpert and bacterial culture, and CSF leucocytes < 5 cells/μL and CSF/blood glucose ratio ≥ 0.5. In Vietnam, patients were included as controls if they had undergone a lumbar puncture, but an alternative, non-infectious, diagnosis was confirmed. In both sites, none of the non-infectious controls received anti-tuberculosis treatment. HIV-negative patients with microbiologically confirmed bacterial meningitis and HIV-positive patients with cryptococcal meningitis patients were included from the same sites.

Ethical approval was obtained from the Ethical Committee of Hasan Sadikin Hospital, Faculty of Medicine, Universitas Padjadjaran, Bandung, Indonesia and from the Oxford Tropical Research Ethics Committee in the United Kingdom, the Institutional Review Boards of the Hospital for Tropical Diseases and Pham Ngoc Thach Hospital in Vietnam. Written (Vietnam) or oral (Indonesia) consent to be included in the study, for storage of surplus sample, and to obtain follow-up data was obtained from patients or close relatives of patients who were unconscious. The paper adheres to the STROBE methodology.

### Metabolite measurements

CSF and blood samples were centrifuged for 15 minutes according to local protocols (865 - 3000 x g) and supernatants were stored at -80’C.^9^ CSF and plasma metabolites were measured using targeted a liquid chromatography tandem mass spectrometry (LC-MS) method with a system comprised of a 1290 Infinity II U-HPLC coupled to an Agilent 6495 Triple Quadrupole mass spectrometer (Agilent Tech. Santa Clara, CA). Metabolites were extracted from plasma or CSF (10 µL) using 90 µL of acetonitrile/methanol/formic acid (74.9:24.9:0.2 v/v/v) containing stable isotope-labeled internal standards (valine-d8, Sigma-Aldrich; St. Louis, MO; and phenylalanine-d8, Cambridge Isotope Laboratories, Andover, MA). The samples were centrifuged (10 min, 9,000 x g, 4°C), and the supernatants were injected directly onto a 150 × 2 mm, 3 µm Atlantis HILIC column (Waters; Milford, MA). The column was eluted isocratically at a flow rate of 250 µL/min with 5% mobile phase A (10 mM ammonium formate and 0.1% formic acid in water) for 0.5 minute followed by a linear gradient to 40% mobile phase B (acetonitrile with 0.1% formic acid) over 10 minutes. Pairs of pooled samples generated using aliquots of all samples in the study were included every 20 samples correct for MS sensitivity drift and for quality control analyses. Sample stability over the 7 years study inclusion and 4-year storage time was checked by plotting metabolite levels of definite TBM patients against storage time. Tryptophan metabolites were measured using the following multiple reaction monitoring transitions: 3-hydroxyanthranilic acid (154.1 -> 136.0), 3-indoleacetic acid (176.1 -> 130.1), 3-methoxyanthranilate (168.1 -> 150.0), 5-hydroxyindoleacetic acid (192.1 -> 146.0), 5-methoxytryptophol (192.1 -> 130.0), kynurenic acid (190.1 -> 144.1), kynurenine (209.1 -> 94.0), tryptophan (205.1 -> 187.9), N-acetylserotonin (219.1 -> 160.0), niacinamide (123.1 -> 80.1), quinolinic acid (168.0 -> 149.9) and xanthurenic acid (206.1 -> 132.0). Absolute concentrations were determined using external calibration curves created via serial dilution of stable isotope-labeled compounds in CSF and plasma. These compounds were sourced from Cambridge Isotope Labs: 3-indoleacetic acid-d7 (DLM-8040), anthranilic acid-^13^C6 (CLM-701), 5-HIAA-^13^C6 (CLM-9936), kynurenic acid-d5 (DLM-7374), L-kynurenine-d6 (<u>DLM-7842</u>), L-tryptophan-^13^C11 (CLM-4290) and niacinamide-^13^C6 (CLM-9925). Peak abundances were manually integrated using the MassHunter software provided by the LC-MS manufacturer.

### CSF mycobacterial load and inflammatory proteins

The CSF mycobacterial load was inferred qualitatively by comparing patients with negative versus positive CSF culture, and semiquantitively from the GeneXpert Ct-values as described previously,^10^ and inferred from CSF *M. tuberculosis* culture. CSF inflammatory cytokines in 178 Indonesian HIV-negative TBM patients were measured using a multiplex proximity extension assay (Olink) in two batches. Olink uses a multiplex assay that simultaneously recognize 96 target proteins through specific paired-antibodies which are coupled with unique oligonucleotides, for quantitative PCR measurement.^11^ For each protein, overlapping samples from two batches were fitted in a linear regression model, where the linear components were subsequently extracted, and used as correction factors for batches normalization. In 304 Vietnamese HIV-negative patients, 10 human cytokines were measured in CSF with Luminex multiplex bead array technology (Bio-Rad Laboratories, Hercules, CA).^4^ CSF total protein was used as proxy for blood-CSF barrier disruption as it showed a near-perfect correlation with the established marker CSF-serum albumin (r^2^=0.98).^12^

### Quality control and statistical analysis

Only metabolites and proteins with a coefficient of variation (CV) of the pooled samples < 30% and < 25% missing values, respectively among TBM patients were further included in the analysis. Remaining missing metabolite values after quality control were replaced with half of the minimum measured value of the corresponding metabolite, and log2-transformed subsequently. Statistical analyses were performed in R 4.0.4^13^, using the R packages survival, tableone, dplyr, openxlsx, pheatmap, grid, and ggplot2. Correlation analyses between metabolites levels, and between metabolites levels and clinical and inflammatory parameters, were calculated using spearman-rank correlation. The impact of baseline CSF and plasma metabolite levels on 60-day survival was tested in a Cox-regression model, adjusted for sex, age, and HIV status as covariates. The model stratified by study site as mortality is known to be higher in the Indonesian^5^ than in the Vietnamese^7^ cohort. An analysis plan was made before the study, and correction for multiple testing using the Benjamini Hochberg method was done if multiple comparisons were done in primary analysis.

## Results

### Baseline characteristics of TBM patients and controls

We studied 1069 adults with TBM, 390 from Indonesia and 679 from Vietnam (**Table 1**). Patients were young (median age 34 years), 26.6% were HIV-positive, and the majority presented with a moderately severe (55.6 % grade II) to severe (17.0% grade III) severe disease according to the international classification.^14^ The rate of mycobacterial confirmation was 64.1%. Sixty-day mortality, the primary endpoint in the analysis, was 21.6%. Patients who died within 180 days from admission did so after a median of 14 days. A 14-day cut-off was therefore used to distinguish early from late mortality as a secondary endpoint.

**Table 1:** Patient baseline characteristics.

|  | Tuberculous meningitis<br>(n=1069) | Non-infectious control<br>(n=54) | Bacterial meningitis<sup>1</sup><br>(n=50) | Cryptococcal meningitis<sup>1</sup><br>(n=60) |
| --- | --- | --- | --- | --- |
| **Clinical features** |  |  |  |  |
| Age, years | 34 (27-44) | 35 (25- 44) | 46 (34- 57) | 33 (27- 37) |
| sex - % male | 700 (65.5%) | 30 (55.6%) | 12 (60.0%) | 26 (78.8%) |
| Glasgow Coma Scale | 14 (12-15) | 15 (12-15) | 13 (9-14) | 15 (13-15) |
| HIV - % positive | 284 (26.6%) | 11 (20.4%) | 0 (0%) | 60 (100%) |
| Tuberculous meningitis grade (%) |  |  |  |  |

**Table 1: Patient baseline characteristics.**
|  |  |  |  |  |
| --- | --- | --- | --- | --- |
| Grade I | 287 (27.3%) | - | - | - |
| Grade II | 584 (55.6%) | - | - | - |
| Grade III | 179 (17.0%) | - | - | - |

| Cerebrospinal fluid features |  |  |  |  |
| --- | --- | --- | --- | --- |
| Leukocytes- cells per μL | 150 (49-336) | 2 (1-3) | 1900 (739-5460) | 86 (24-192) |
| Neutrophils- cells per μL | 22 (3-99) | 1 (0-1) | 1527 (538-4986) | 17 (6-109) |
| Mononuclear cells- cells per μL | 98 (38-207) | 2 (1-3) | 307 (134-646) | 31 (6-89) |
| Protein- g/L | 1.46 (0.90-2.40) | 0.40 (0.26-0.59) | 1.90 (1.10-3.80) | 0.76 (0.58-1.60) |
| CSF to blood glucose ratio | 0.28 (0.17-0.40) | 0.60 (0.56-0.70) | 0.46 (0.17-1.00) | 0.50 (0.30-1.00] |
| *M. tuberculosis* culture or ZN-staining or GeneXpert positive | 686 (64.17%) | - | - | - |

| Outcomes |  |  |  |  |
| --- | --- | --- | --- | --- |
| Outcome at day 60 |  |  |  |  |
| Alive | 825 (77.2%) | - | - | - |
| Deceased | 231 (21.6%) | - | - | - |
| Lost to follow-up | 13 (1.2%) | - | - | - |
| Outcome at day 180 |  |  |  |  |
| Alive | 731 (68.4%) | - | - | - |
| Deceased | 304 (28.4%) | - | - | - |
| Lost to follow-up | 34 (3.2%) | - | - | - |

Legend: categorical variables are presented in N (%); Continuous variables are presented in median (IQR). Abbreviations: CSF=cerebrospinal fluid. <sup>1</sup>Clinical metadata available for 40% of bacterial meningitis and 56% of cryptococcal meningitis patients.

There were some differences between the populations. Indonesian patients presented with more severe diseases (91.9% grade with grade II or III) than Vietnamese patients (62.2%). Also, CSF total protein, a proxy for blood-CSF barrier leakage,^12^ was higher in Indonesian (median=1.6 g/L, IQR=0.8-3.1) than Vietnamese (1.3 g/L, IQR=0.8-2.0) patients. CSF polymorphonuclear cell counts were higher in the Indonesian than in the Vietnamese patients where it showed a bimodal distribution associated to study site **(Supplementary Figure 1)** and stratified analyses were performed taking this into account. Compared to the TBM patients, non-infectious controls (n=54), bacterial meningitis patients (n=50), and cryptococcal meningitis patients (n=60) had a similar age range and gender distribution.

Ten metabolites showed detectable levels in >75% of patients and passed quality control, while two metabolites, 3-methoxyanthranilate and 5-hydroxyindoleacetic acid, were detected in less than 75% of patients and excluded from further analysis. Metabolite measurements showed stable concentrations over and were not affected by year of patient inclusion and duration of sample (**Supplementary Figure 2**).

### Increased CSF tryptophan levels were associated with mortality of TBM patients independent of HIV status

Confirming our previous findings,^3^ higher baseline CSF tryptophan levels predicted 60-day survival in patients with TBM (HR=1.16 for each doubling in CSF tryptophan, 95%CI=1.10-1.24), all analyses corrected for age, sex and HIV status, and stratified for cohort, **Figure 2, Table 2**). This was both true for HIV-negative (HR=1.13, 95%CI=1.05-1.21) and HIV-positive patients (HR=1.19, 95%, CI=1.07-1.33), who showed a much higher mortality **(Supplementary Figure 3)**, as reported previously.^4,5^ Baseline CSF tryptophan was associated with both early (HR=1.14, 95%CI=1.06-1.23) and late (HR=1.17, 95%CI=1.08-1.26) mortality (**Supplementary Table 1)**. Compared to non-infectious controls, CSF tryptophan was lower. This was also observed in patients with cryptococcal, but not in those with bacterial meningitis (**Figure 3**). Interestingly, among 17 cryptococcal meningitis patients with available in-hospital mortality data in Indonesia, baseline CSF tryptophan was significantly higher in those who died in hospital compared to those discharged alive (**Supplementary Figure 4**), similar as in TBM.

**Table 2.** Univariate Cox regression for 60-day mortality for CSF metabolites.

| Metabolites | HR<sup>1</sup> | 95% CI<sup>1</sup> | p-value | FDR<sup>2</sup> |
| --- | --- | --- | --- | --- |
| tryptophan | 1.16 | 1.10, 1.24 | <0.001 | **<0.001** |
| kynurenine | 1.00 | 0.93, 1.07 | >0.9 | >0.9 |
| kynurenic acid | 1.00 | 0.93, 1.07 | 0.9 | >0.9 |
| 3-hydroxyanthranilic acid | 1.01 | 0.97, 1.05 | 0.6 | 0.9 |
| xanthurenic acid | 0.95 | 0.90, 1.00 | 0.05 | 0.2 |
| quinolinic acid | 0.92 | 0.85, 1.00 | 0.038 | 0.2 |
| niacinamide | 1.03 | 0.95, 1.11 | 0.5 | 0.8 |
| 3-indoleacetic acid | 1.11 | 0.96, 1.29 | 0.2 | 0.4 |
| N-acetylserotonin | 1.01 | 0.94, 1.09 | 0.7 | 0.9 |
| 5-methoxytryptophol | 1.11 | 0.93, 1.32 | 0.3 | 0.5 |

Legend: Cox regression models were stratified by cohort and adjusted by sex, age, and HIV status. Hazard ratio (HR) was calculated per 2-fold increase in metabolite concentration. <sup>1</sup> HR = Hazard ratio, CI = Confidence interval. <sup>2</sup> FDR=False Discovery Rate; Benjamini & Hochberg correction for multiple testing.

**Figure 2.**
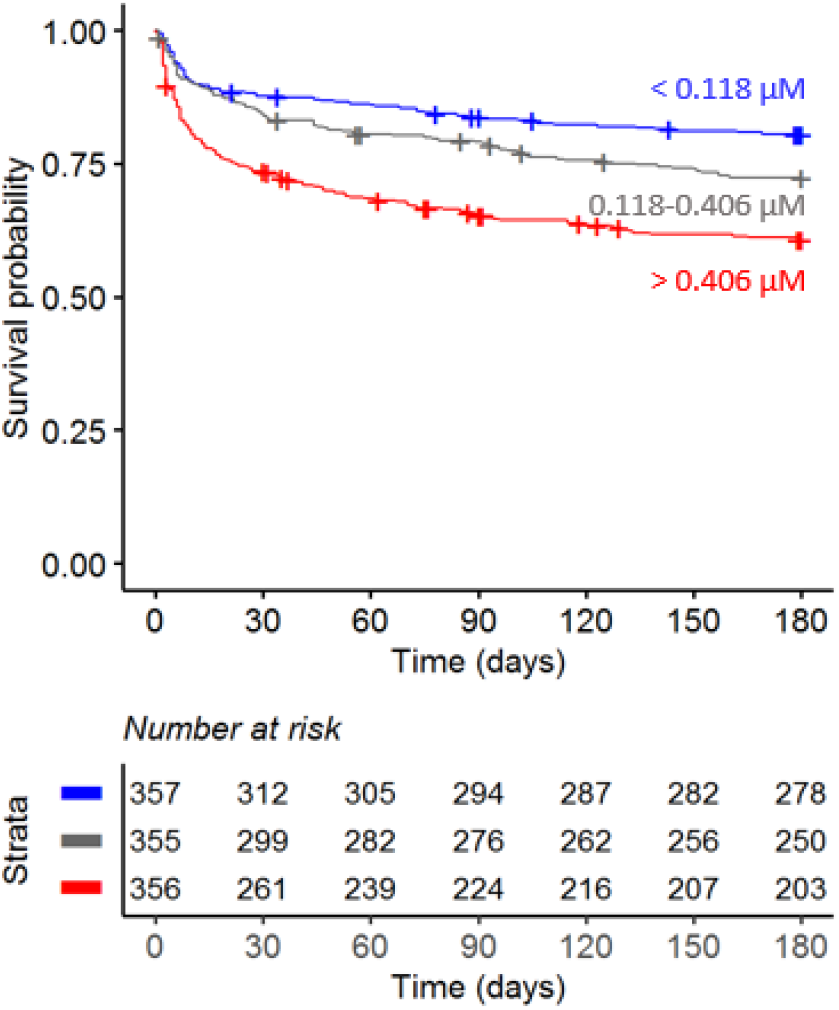
Six-months survival curve of TBM patients. Patients were stratified by CSF tryptophan concentrations tertiles.

**Figure 3.**
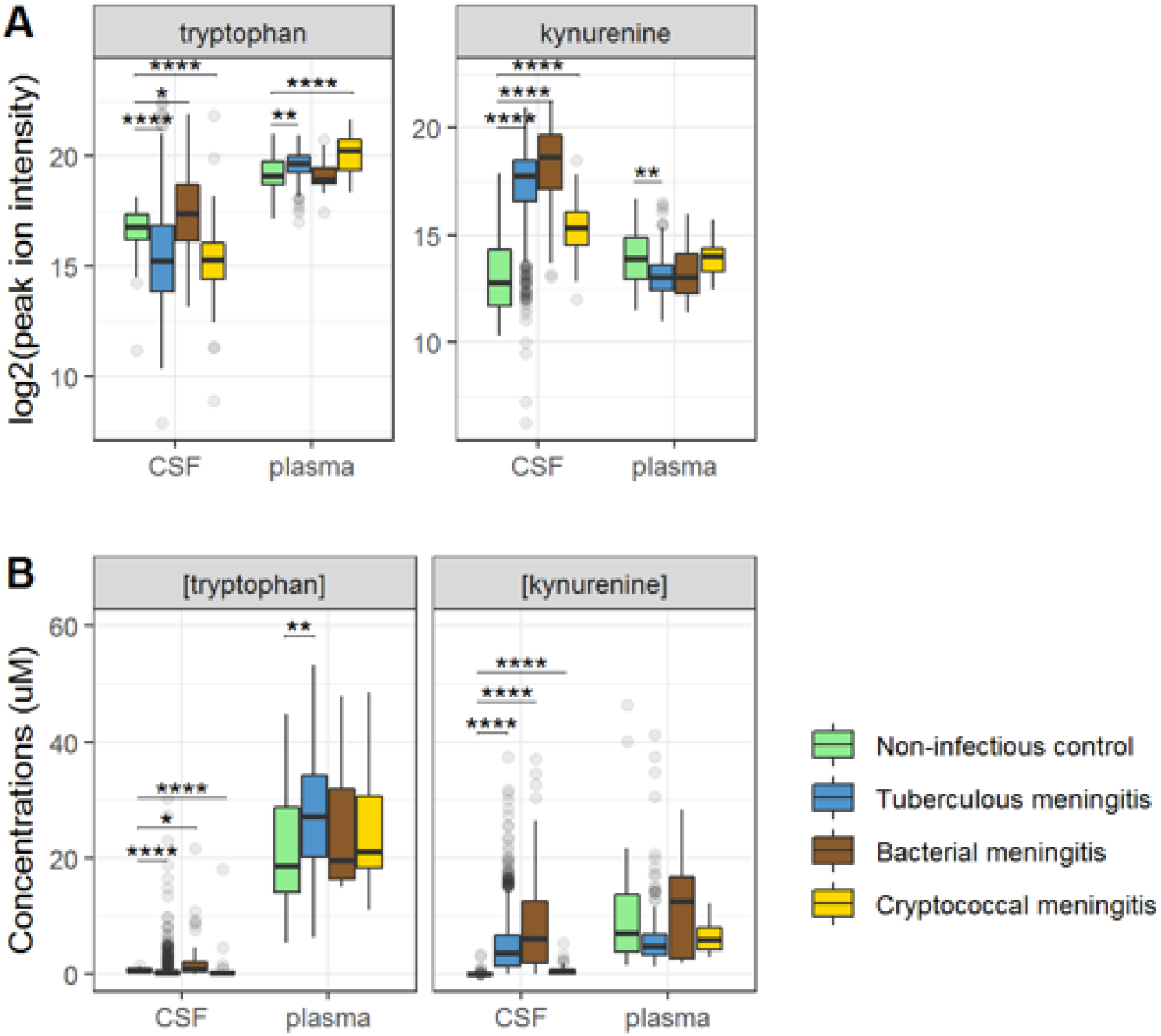
CSF and plasma metabolites concentrations in TBM and controls for the tryptophan metabolites associated with outcome: tryptophan and kynurenine. (A) Relative concentrations based on peak ion intensity (B) absolute concentrations in μM.

### CSF tryptophan levels do not reflect mycobacterial burden

We next examined if CSF tryptophan was associated with CSF mycobacterial load. We hypothesized that a low baseline tryptophan might either reflect a lower bacterial load, as *M. tuberculosis* can produce tryptophan, *or* might cause a lower bacterial load as tryptophan depletion impairs mycobacterial growth.^79^ Instead, we found a reverse, albeit weak relationship: tryptophan was higher in CSF culture negative (median=0.31 µM) than culture positive (median=0.14 µM, p<0.001) TBM patients. Similarly, among patients with CSF GeneXpert-confirmed tuberculous meningitis patients, we did not find a correlation between CSF tryptophan and quantitative PCR results (Spearman’s rho=0.084, p=0.105, **Supplementary Figure 5)**. Interestingly, within patients with microbiologically confirmed TBM the effect of tryptophan was stronger (HR=1.28, 95%CI=1.17-1.40) than in patients with probable TBM (HR=1.07, 95%CI=0.98-1.18).

### Relationship between cerebral and systemic metabolism and its impact on survival

Ninety-five percent of tryptophan is converted to kynurenine^6^ and we therefore hypothesized that lower CSF tryptophan levels in TBM are caused by higher conversion to kynurenine, and that the higher CSF tryptophan associated with death could reflect reduced activity of IDO1 and other downstream enzymes. CSF kynurenine (**Figure 3**) and its downstream metabolite kynurenic acid (**Supplementary Figure 6**) were higher in TBM patients, bacterial meningitis and cryptococcal meningitis patients compared to non-infectious controls, but not significantly different between surviving and non-surviving TBM patients (**Table 2**).

Then, to examine the relation between cerebral and systemic tryptophan metabolism, we compared concentrations of CSF metabolites with those in plasma, measured in a subset of 300 TBM patients. In contrast to our findings in CSF, plasma tryptophan levels were higher and kynurenine levels were lower in TBM patients compared to controls. As the CSF kynurenine metabolites positively correlated with CSF protein (**Figure 4**), a proxy for barrier leakage^12^, we hypothesized that systemic leakage might be an additional source of kynurenine. For a subset of metabolites, absolute quantification of metabolite levels was achieved. This showed that the increase in CSF kynurenine in TBM patients (Δ = 3.52 µM) was much more marked than the decrease in CSF tryptophan (Δ = 0.39 µM, **Figure 3B**). Corroborating our leakage hypothesis, the CSF-plasma gradient of the kynurenine metabolites showed strong correlations with total CSF protein (**Supplementary Figure 7**). Plasma tryptophan did not predict mortality, but plasma levels of its downstream metabolites kynurenine strongly predicted mortality (**Supplementary Table 2)**.

**Figure 4.**
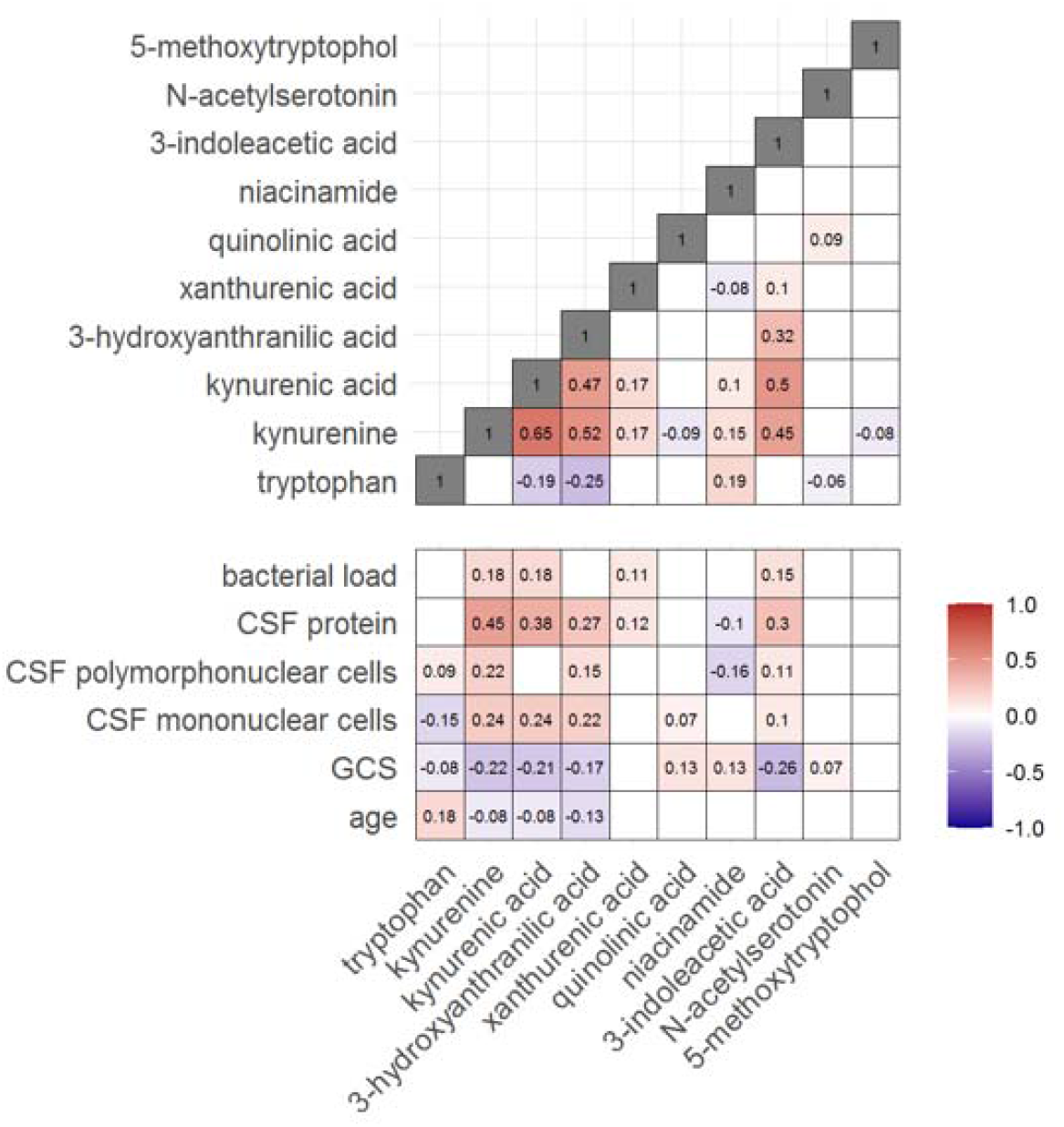
Correlation between tryptophan metabolites and with clinical and CSF parameters. Significant Spearman’s correlation coefficients are presented in the correlation matrix, while the ones with not significant correlations were blank. Red indicates positive correlations, and blue indicates negative ones. The color gradient shows the strength of the associations.

### CSF tryptophan is inversely correlated with interferon-gamma

We next looked at correlations of tryptophan metabolites and inflammation, as inflammation is a determinant of outcome from TBM.^2^ Out of 92 inflammatory proteins measured in CSF from 176 TBM patients from Indonesia, 80 proteins were detectable in >75% of patients. Tryptophan correlated inversely to a small cluster of 13 cytokines, including interferon gamma (IFN-γ, **Supplementary Figure 8**). In line with this finding, a higher CSF IFN-γ has previously been shown to predict survival of Vietnamese TBM patients.^4^ IFN-γ is known to induce IDO1^15^, which converts tryptophan to kynurenine. We indeed confirmed the inverse correlation between CSF tryptophan and IFN-γ in our Vietnamese patients (Spearman’s rho=-0.45, p<0.0001, **Figure 5A**), irrespective of HIV-status. Different from tryptophan, the kynurenine metabolites (kynurenine, kynurenic acid, 3-hydroxyanthranilic acid, and quinolinic acid) correlated strongly with a large cluster of inflammatory proteins including the hallmark inflammatory protein TNF-α, which we could again confirm in the Vietnamese patients (Spearman’s rho=0.30, p<0.0001, **Figure 5B**).

**Figure 5.**
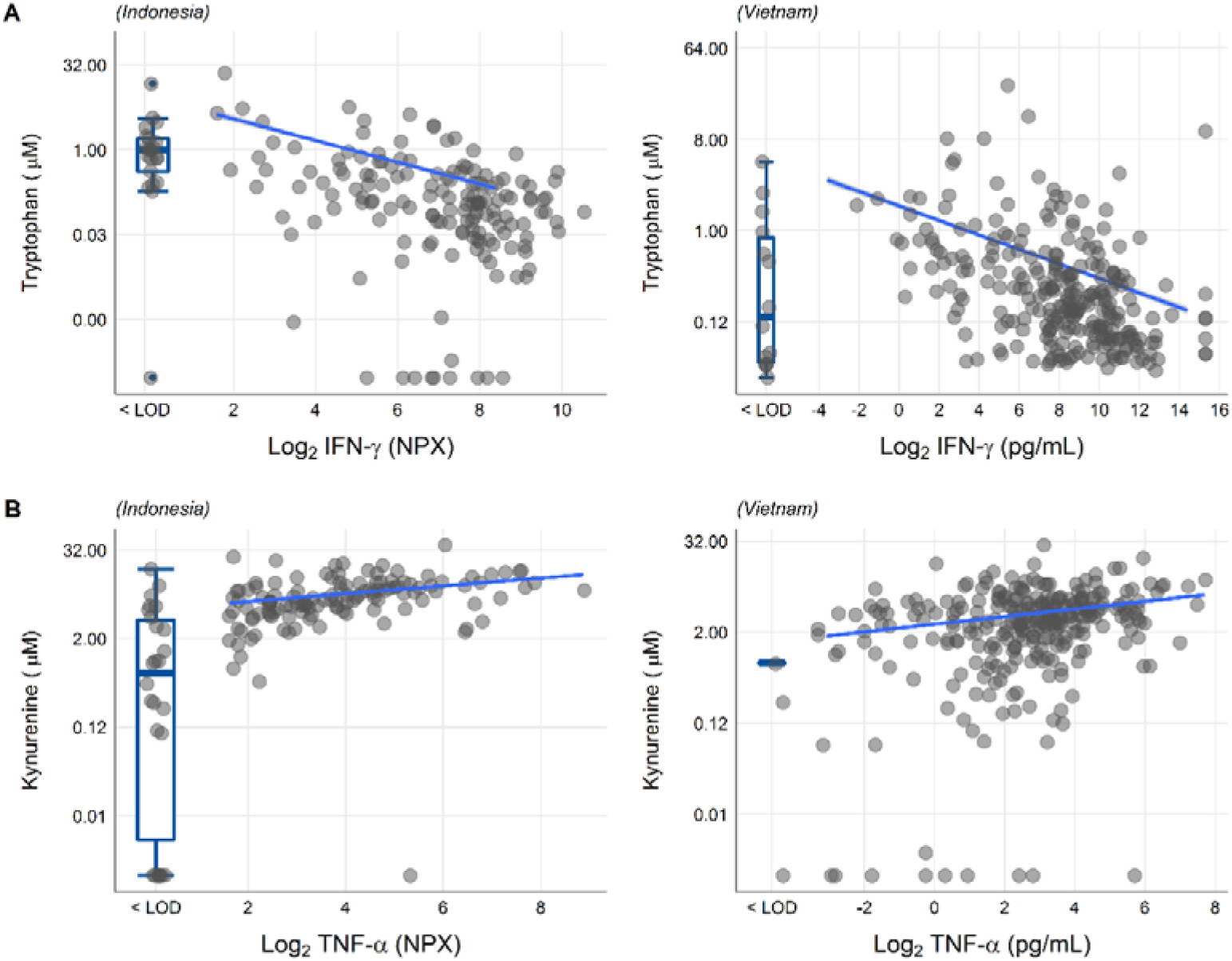
Associations of CSF tryptophan with IFN-γ (A) and with TNF-α (B) in 176 Indonesian (left) and 304 (Vietnamese) TBM patients. The boxplots on the left of each plot show the association of metabolites with cytokines below the detection limit. Abbreviations: IFN-γ: interferon gamma, TNF-α: tumor necrosis factor alpha, LOD: lower limit of detection.

## Discussion

We previously found that CSF concentrations of tryptophan were lower in HIV-negative Indonesian adults with TBM compared to non-infectious controls, and that TBM patients with lower tryptophan levels had lower mortality.^3^ In the current study we confirm these observations in a much larger cohort of HIV-negative and positive patients from both Vietnam and Indonesia. Aiming to understand how tryptophan metabolism is altered in TBM and how it might exert its effect on patient outcome, we correlated its concentrations with bacterial load and CSF inflammatory markers and measured downstream metabolites both in CSF and plasma. Our findings show that CSF concentrations of downstream kynurenine metabolites did not predict mortality, and that higher tryptophan levels were not associated with a higher bacterial load. Also, while kynurenine metabolites strongly correlated with CSF inflammatory markers and CSF protein, a marker of blood-CSF leakage, there was no association with CSF tryptophan. A higher tryptophan did however show a strong negative correlation with IFN-γ, important for immunity against mycobacteria. Collectively, these findings suggest that tryptophan affects outcome from TBM within the brain rather than systemically. This is potentially driven by IFN-γ but not associated with nonspecific inflammation, and independent from downstream tryptophan metabolism or bacterial replication. In contrast, kynurenine may affect outcome systemically by leakage across the blood-brain barrier.

CSF tryptophan increases with age in individuals without central nervous system infections.^16^ Age is known to negatively impact outcome of TBM^4,5^ and in this study, higher age was associated with higher CSF tryptophan concentrations. All mortality analyses were therefore corrected for age, as well as sex and HIV-status, and analysis was stratified for country because of the overall higher mortality in Indonesian compared to Vietnamese tuberculous meningitis patients.^4,5^ We further tested whether higher CSF tryptophan reflected a higher mycobacterial burden and refuted this hypothesis. For cryptococcal meningitis, no previous data on cerebral tryptophan metabolism was known. These patients follow a pattern similar to TBM, with low tryptophan and high kynurenine, and in a small number of cryptococcal meningitis patients, a high baseline CSF tryptophan predicted mortality, similar ss for TBM.

Systemic tryptophan and kynurenine are transported into the brain over the large amino acid transporter *LAT1*. In a healthy brain, systemic and CSF kynurenine positively correlate, as do CSF concentrations of tryptophan and kynurenine.^16^ In patients with cerebral inflammation, the correlation between CSF kynurenine and tryptophan can be lost, probably through increased catabolism through IDO upregulation, which also has been demonstrated in the brain parenchyma of deceased TBM patients.^18^ Although we found low CSF tryptophan and high CSF kynurenine in TBM compared to healthy controls, the two did not intercorrelate and moreover, the increase in CSF kynurenine was much larger than the decrease in CSF tryptophan and it is therefore unlikely that upregulation of IDO1 solely explains this which precludes catabolism as the sole explanation. This suggests that increased blood to central nervous system transport as an additional mechanism to IDO1 upregulation. Endothelial cells and pericytes of the blood-brain-barrier can upregulate tryptophan catabolism into kynurenine metabolites upon IFN-γ stimulation.^19^ Our findings corroborate this hypothesis because we find a strong negative correlation between CSF IFN-γ and CSF tryptophan in our patients.

We examined whether higher CSF tryptophan concentrations reflected higher concentrations of downstream kynurenine metabolites that may have neurotoxic (quinolinic acid) or lower levels of the metabolites that may have neuroprotective (kynurenic acid) properties^6^ and refuted these hypotheses. Interestingly however, CSF kynurenine metabolites correlated with CSF cell counts and pro-inflammatory proteins, including TNF-α. Kynurenine is sensed by the aryl hydrocarbon receptor (AhR), which is important for the upregulation of TNF among other pro-inflammatory cytokines in a mouse model,^20^ in line with our CSF findings. The increased CSF kynurenine levels we found in TBM have been reported before in bacterial meningitis^21,22^ and in cerebral malaria^23^ and in plasma from pulmonary TB patients.^17^ Of interest, nicotinamide can inhibit *M. tuberculosis* growth, and can compete with isoniazid for antimycobacterial effects.^24^ We did however not find a detrimental effect of a higher nicotinamide, possibly because of its complex biology, i.e. it can also be produced by *M. tuberculosis* when human dietary niacin intake is limited.^25^

Strengths of our study include the large numbers of clinically well-phenotyped patients from multiple independent study sites in Indonesia and Vietnam, including a significant proportion of HIV-positive patients. We moreover used a sensitive triple quadrupole (QQQ) mass spectrometry method specifically designed to accurately target the tryptophan metabolites. Absolute quantification of a subset of metabolites further facilitated interpretation. Due to differences in polarity of the downstream tryptophan metabolites, we could not measure the complete tryptophan pathway. The availability of CSF at baseline only, limits our ability to understand how changes in tryptophan metabolism influence mortality. And we infer our observations from lumbar CSF, which reflects biological processes from both the blood and the brain. The use of ventricular CSF and potentially animal models, could help establishing what processes in the brain parenchyma take place. Animal studies are warranted to see whether pharmacological induction of IDO1 (for instance with recombinant IFN-γ), or inhibitory tryptophan analogues^26^ should be priorities as adjuvant therapeutic candidates for future personalized trials.

In summary, we confirm the importance of CSF tryptophan to outcome from HIV-negative and HIV-positive adults with TBM, independent from downstream kynurenine metabolism, bacterial load and inflammation. We additionally show the potential importance of systemic kynurenine as a predictor of mortality. Better understanding of the metabolic pathways associated with TBM may lead to more targeted therapies, as adjuvant immunotherapy may modulate the aberrant metabolic pathways and thus improve outcome.

## Data Availability

The data used in this manuscript will be available as supplementary material.

## Materials availability

The clinical metadata and LC-MS data before pre-processing is available as Supplementary File to this manuscript.

## Acknowledgements

The authors thank the neurology residents and Tiara Pramaesya, Sofia Immaculata, Putri Andini, Sri Margi, Rani Trisnawati and Shehika Shulda of the tuberculous meningitis study team for monitoring patients and data management; Lidya Chaidir and Jessi Annisa for mycobacterial diagnostics; the director of the Hasan Sadikin General Hospital, Bandung, Indonesia, for accommodating the research. We also express our gratitude to our funders: This study was supported by National Institutes of Health (R01AI145781), the Wellcome Trust (110179/Z/15/Z and 206724/Z/17/Z). Previous establishment of the cohorts in Indonesia was supported by the Direktorat Jendral Pendidikan Tinggi (BPPLN fellowship to SD) and the Ministry of Research, Technology, and Higher Education, Indonesia (PKSLN grant to THA, RR, and SD), and United States Agency for International Development (PEER Health grant to RR). The funders had no role in study design, data collection and analysis, decision to publish, or preparation of the manuscript.

## Supplementary

**Supplementary Figure 1.**
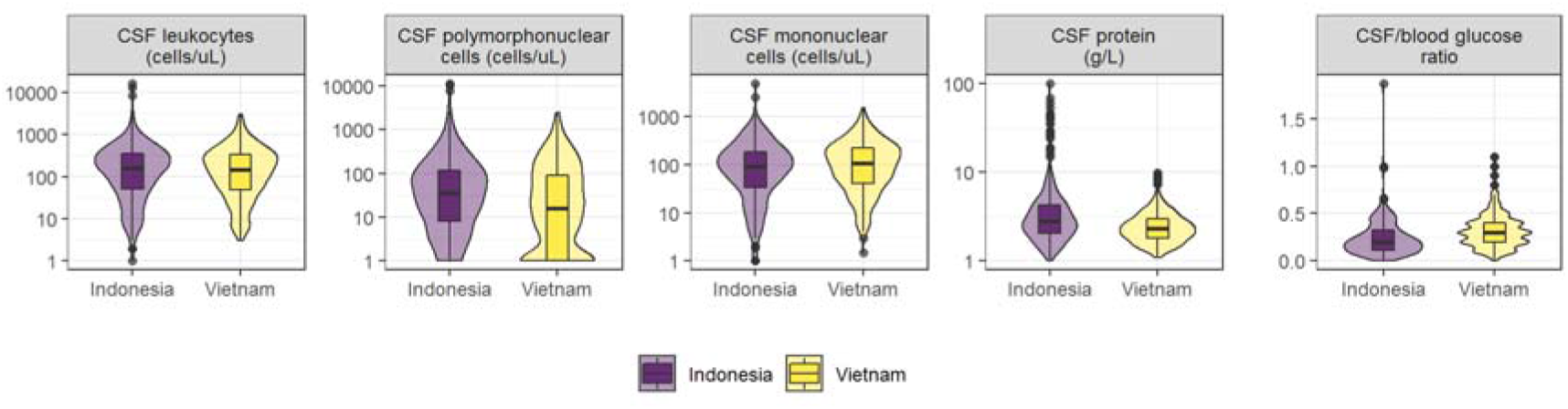
CSF parameters of TBM patients in Indonesia and Vietnam. Distributions of leukocytes, polymorphonuclear cells, mononuclear cells, protein, and the ratio of CSF/blood glucose in Indonesian (purple) and Vietnamese (yellow) patients are depicted by violin plots.

**Supplementary Figure 2.**
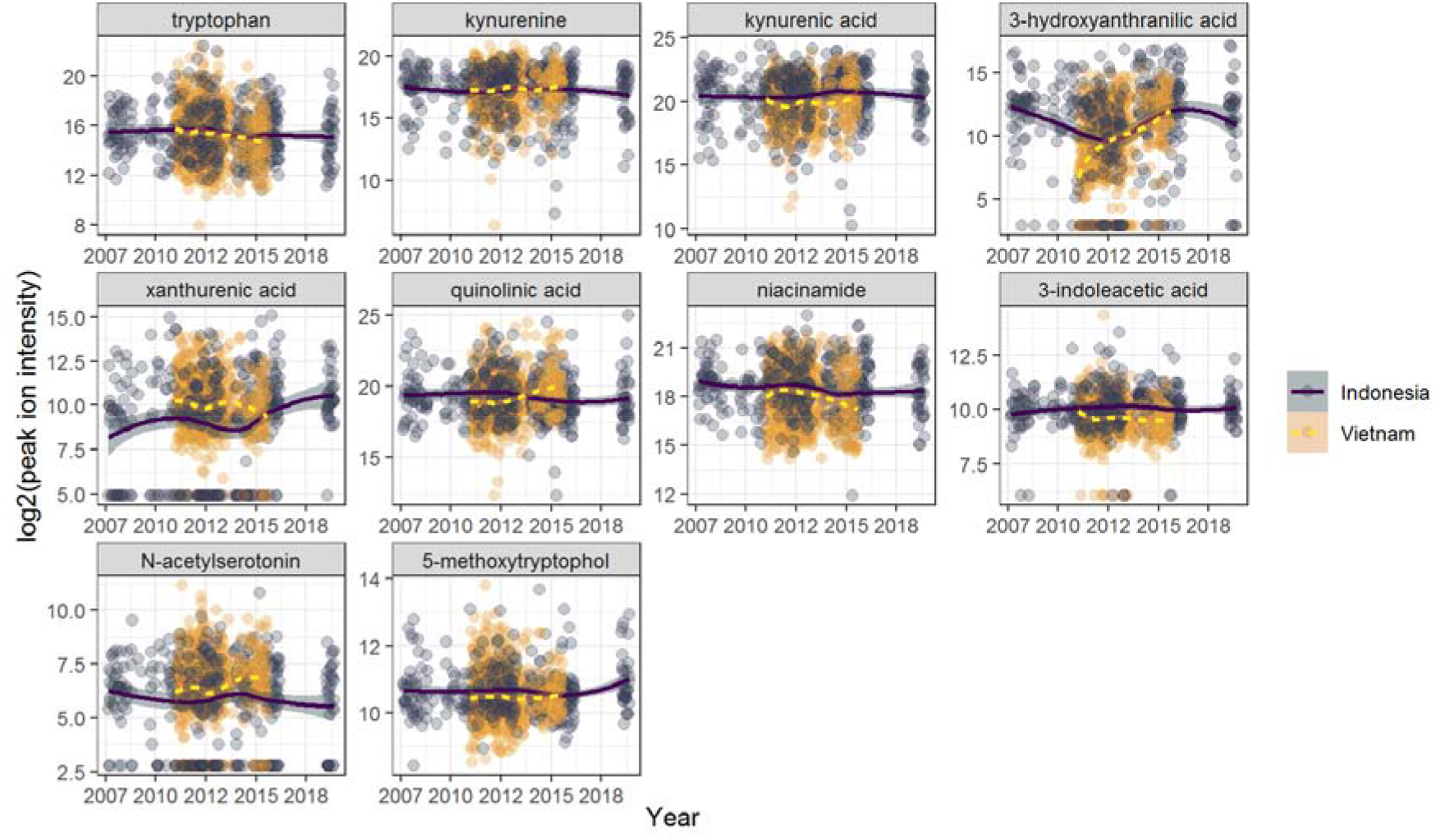
Stability of metabolites over-time. The concentrations of tryptophan metabolites (in log2 scale) were measured in CSF samples from Indonesian (purple) and Vietnamese (yellow) TBM patients were recruited between 2007-2018.

**Supplementary Figure 3.**
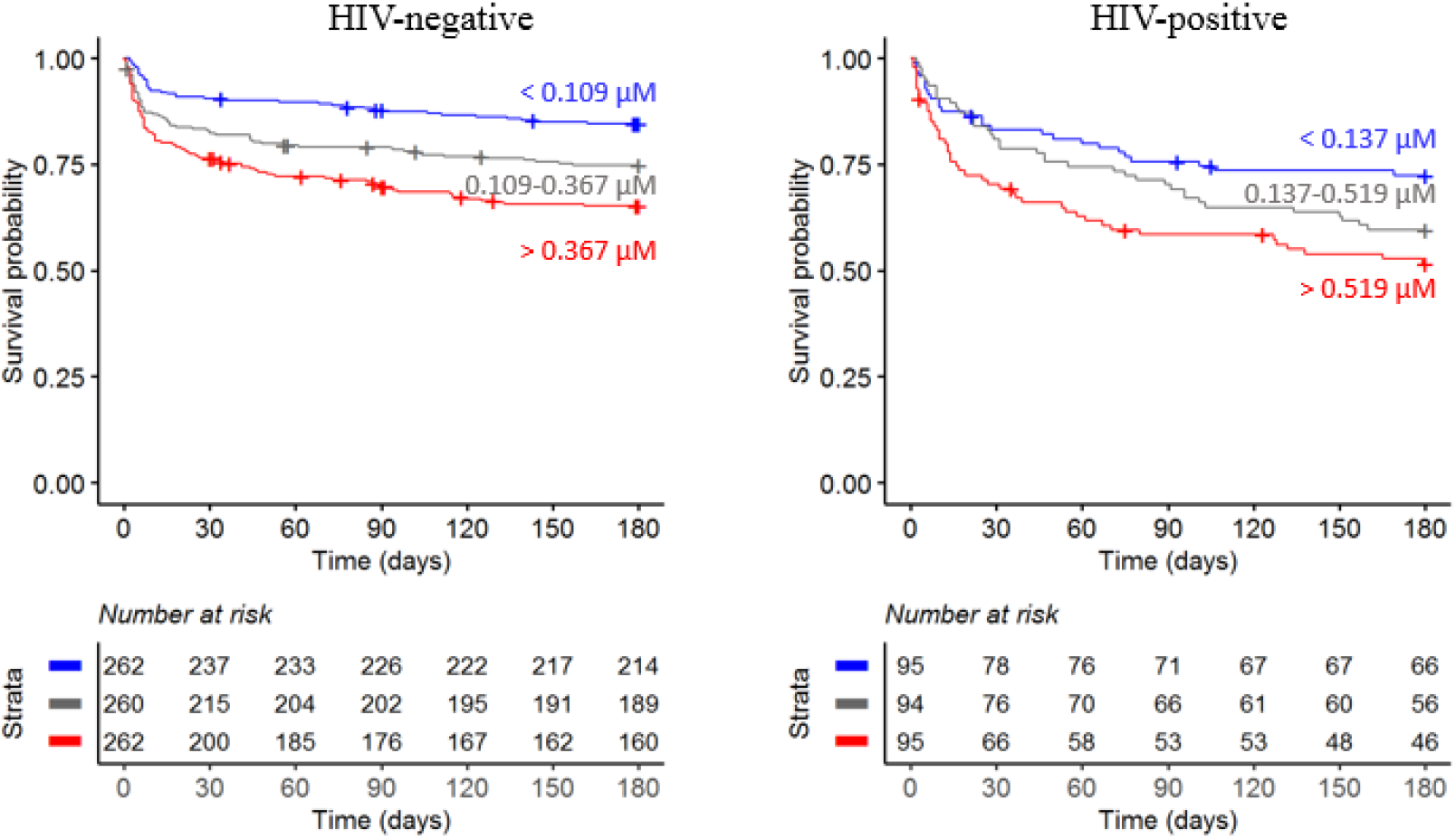
Six-month survival curve of TBM patients stratified by HIV status. Patients were stratified by tertiles based on CSF tryptophan concentrations (red=high tryptophan, gray=intermediate tryptophan, blue=low tryptophan)

**Supplementary Figure 4.**
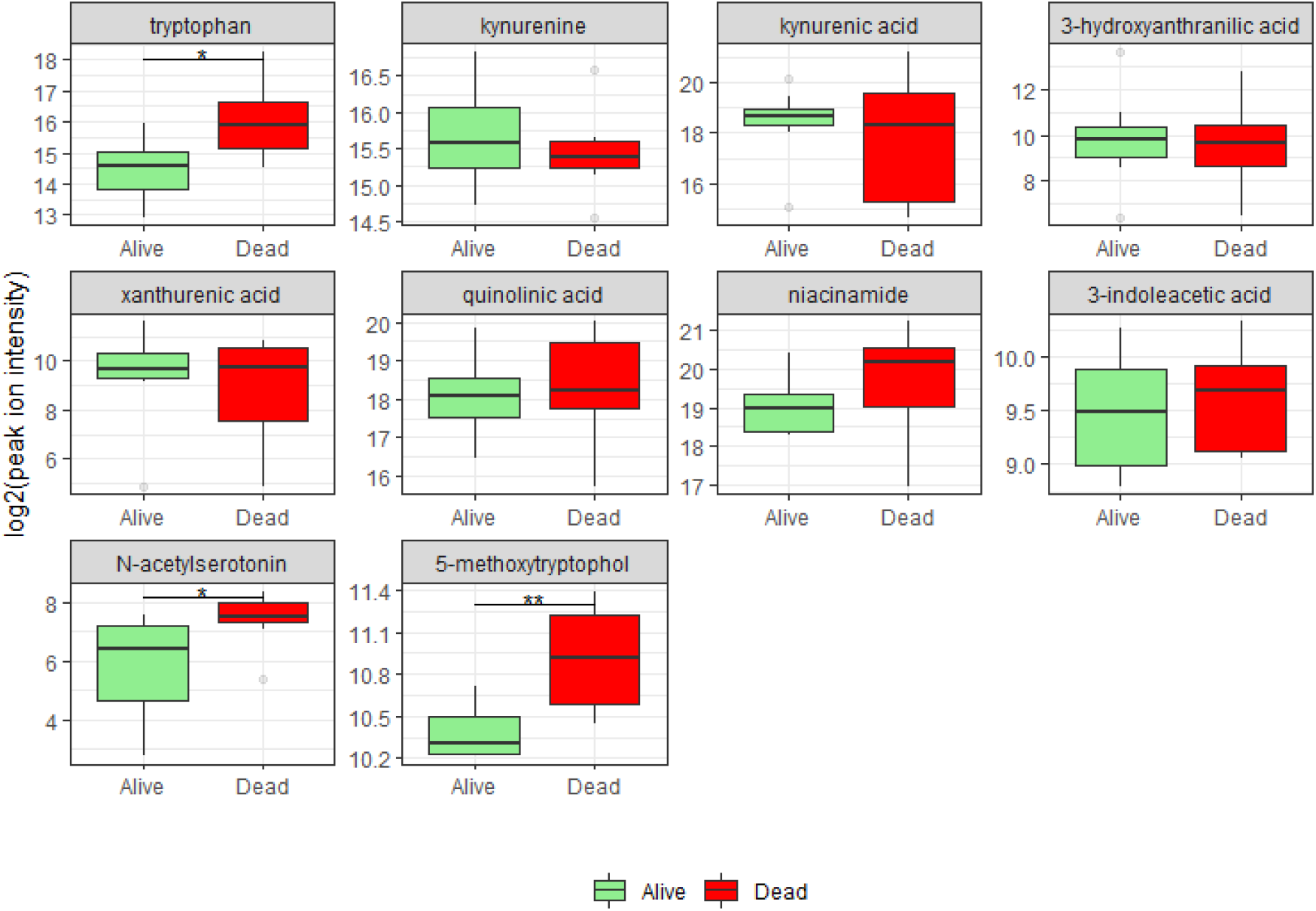
In-hospital mortality for 17 HIV-positive patients with cryptococcal meningitis.

**Supplementary Figure 5.**
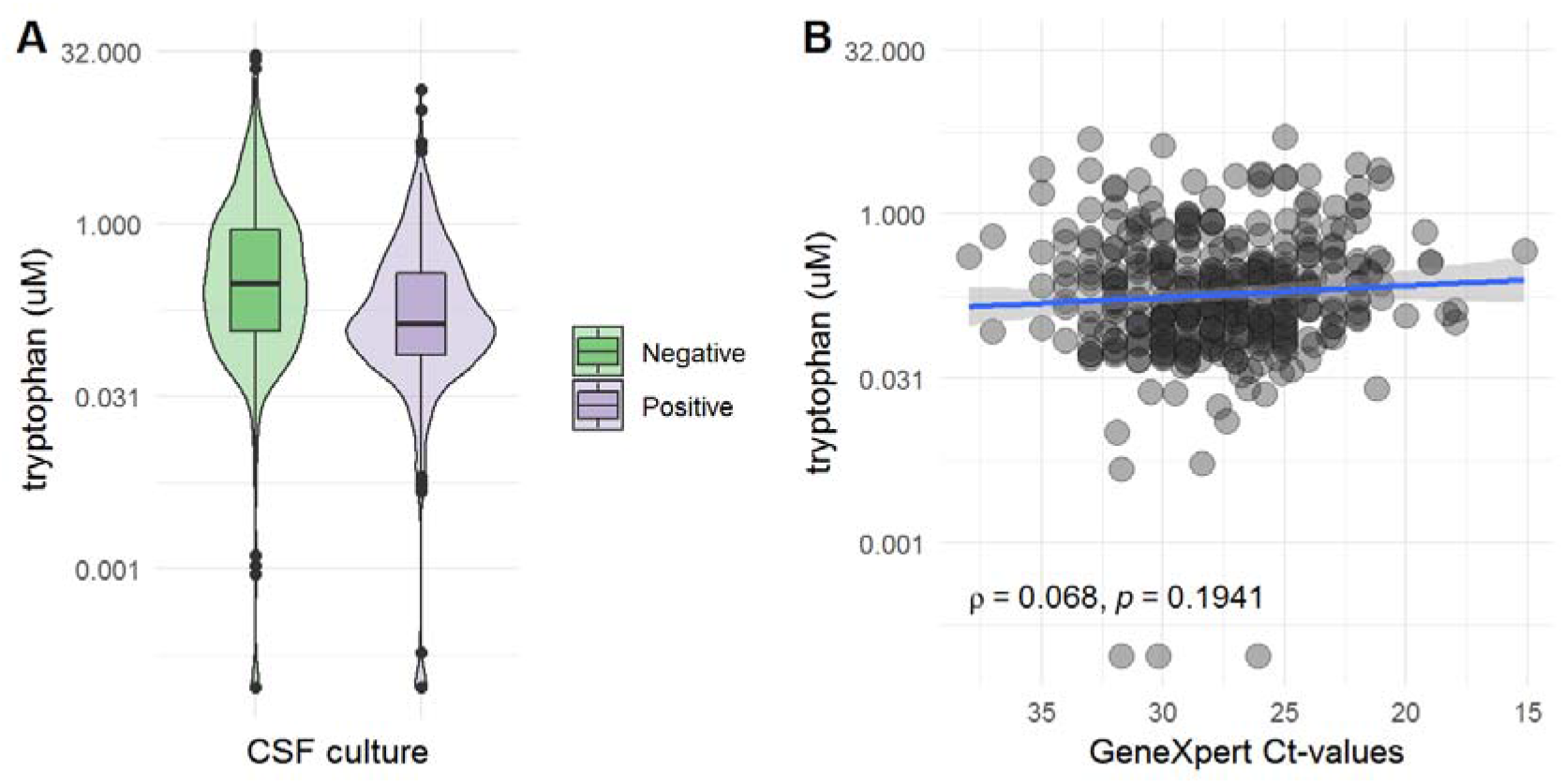
CSF tryptophan distributions according to mycobacterial load. (A) comparing CSF culture negative versus positive patients and (B) among patients with a positive CSF Xpert, in culture positive and culture negative TBM patients, CSF tryptophan was associated with CSF Xpert Ct-values from a low (high Ct-value) to low (low Ct-value) load.

**Supplementary Figure 6.**
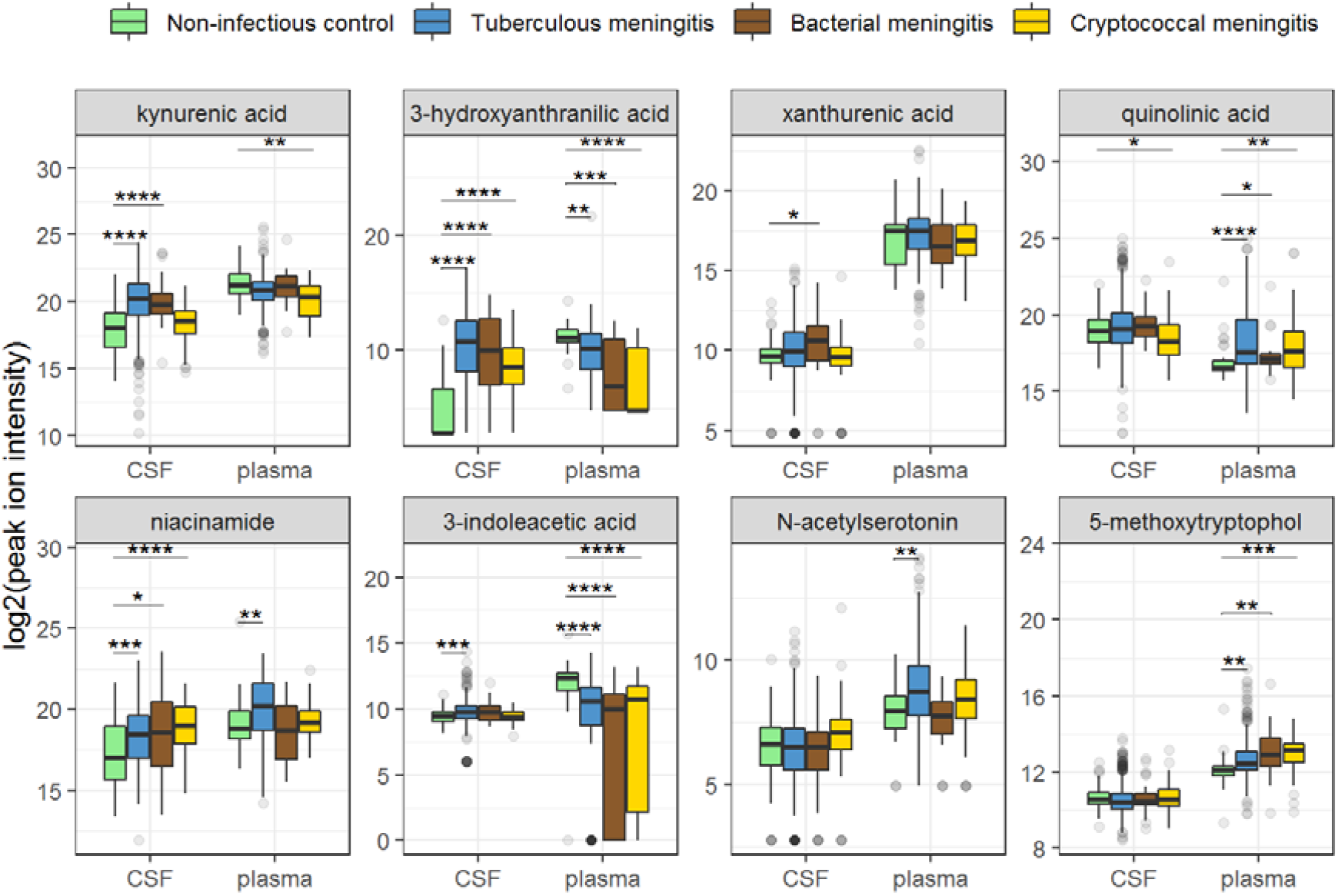
Boxplots of CSF and plasma metabolites concentrations in TBM and controls. Relative concentrations based on peak ion intensities are shown. CSF and plasma concentrations are not directly comparable.

**Supplementary Figure 7.**
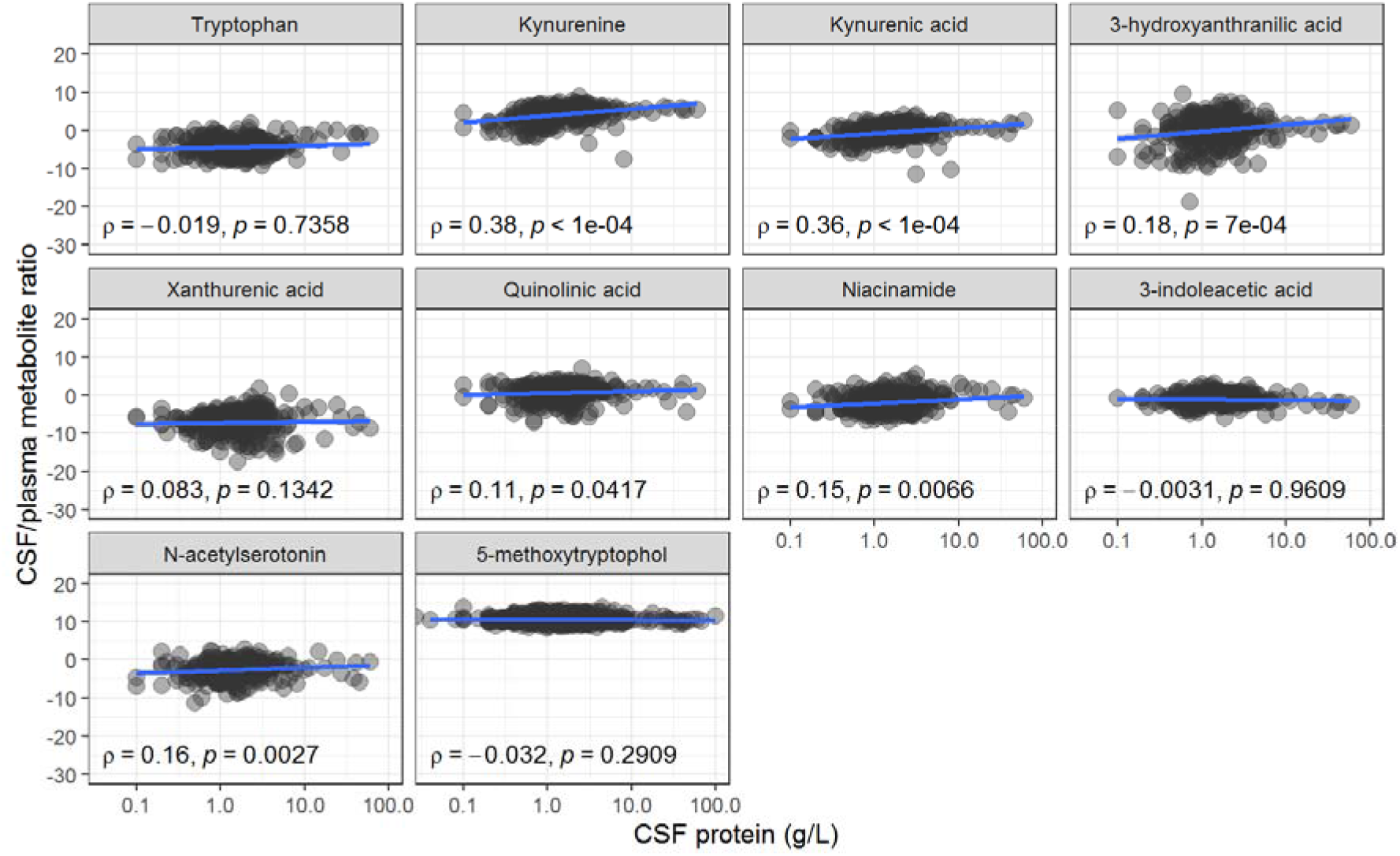
Associations between CSF/plasma metabolite ratios (y-axis) and CSF protein levels (as a proxy of CSF barrier leakage, x-axis). Of note, 71 patients had undetectable plasma levels of 3-indolacetic acid and were removed from this graph.

**Supplementary Figure 8.**
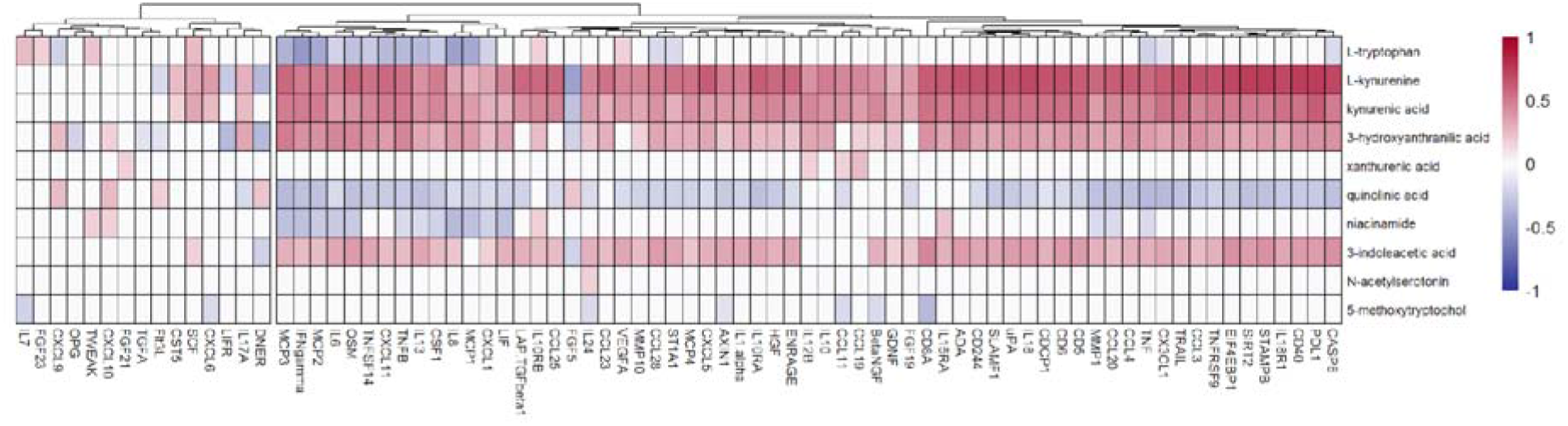
Correlation between CSF tryptophan metabolites and inflammatory markers measured with O-link. Inflammatory markers were clustered based on their correlation coefficients using hierarchical clustering. Red indicates positive correlation, and blue indicated negative correlation.

**Supplementary Table 1.**
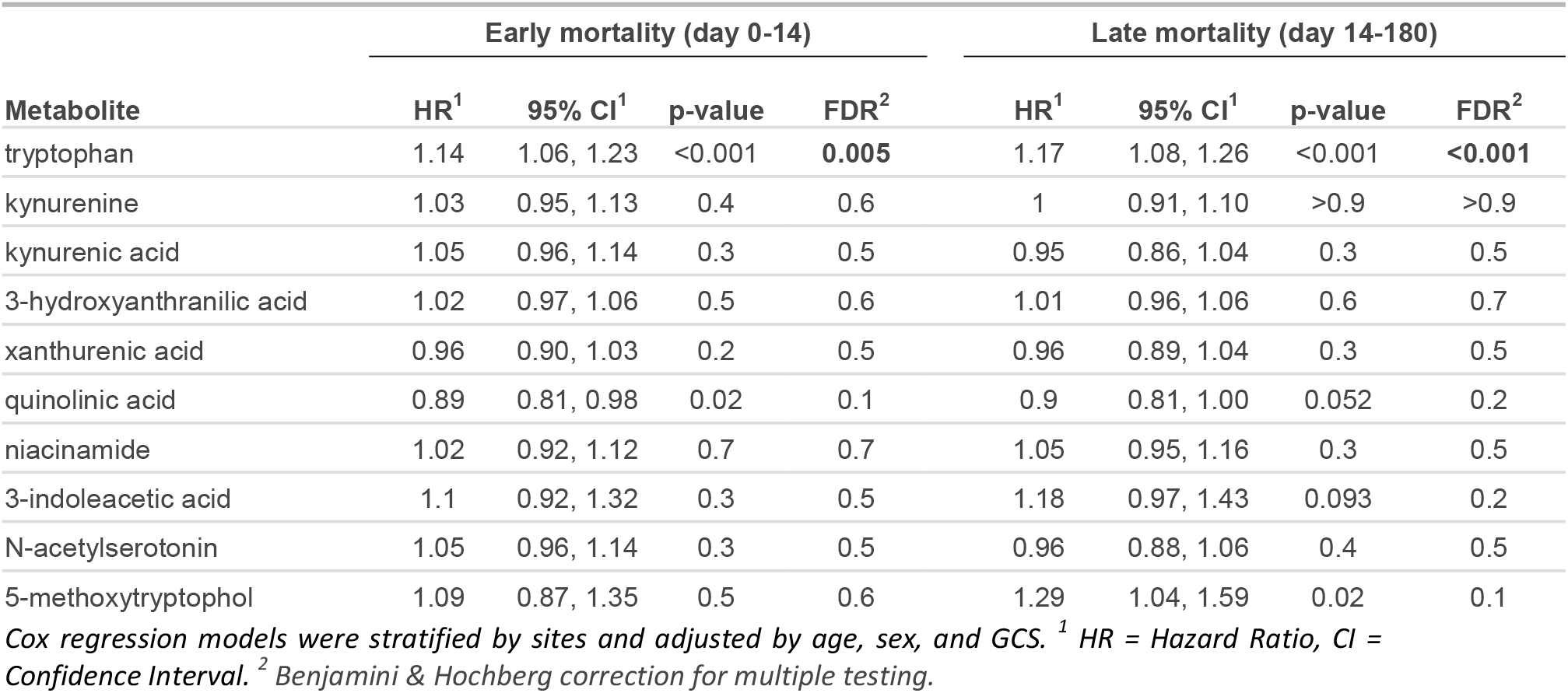
Univariate Cox regression for influence CSF metabolites on early and late mortality.

| Metabolite | Early mortality (day 0-14) |  |  |  | Late mortality (day 14-180) |  |  |  |
| --- | --- | --- | --- | --- | --- | --- | --- | --- |
|  | HR<sup>1</sup> | 95% CI<sup>1</sup> | p-value | FDR<sup>2</sup> | HR<sup>1</sup> | 95% CI<sup>1</sup> | p-value | FDR<sup>2</sup> |
| tryptophan | 1.14 | 1.06, 1.23 | <0.001 | **0.005** | 1.17 | 1.08, 1.26 | <0.001 | **<0.001** |
| kynurenine | 1.03 | 0.95, 1.13 | 0.4 | 0.6 | 1 | 0.91, 1.10 | >0.9 | >0.9 |
| kynurenic acid | 1.05 | 0.96, 1.14 | 0.3 | 0.5 | 0.95 | 0.86, 1.04 | 0.3 | 0.5 |
| 3-hydroxyanthranilic acid | 1.02 | 0.97, 1.06 | 0.5 | 0.6 | 1.01 | 0.96, 1.06 | 0.6 | 0.7 |
| xanthurenic acid | 0.96 | 0.90, 1.03 | 0.2 | 0.5 | 0.96 | 0.89, 1.04 | 0.3 | 0.5 |
| quinolinic acid | 0.89 | 0.81, 0.98 | 0.02 | 0.1 | 0.9 | 0.81, 1.00 | 0.052 | 0.2 |
| niacinamide | 1.02 | 0.92, 1.12 | 0.7 | 0.7 | 1.05 | 0.95, 1.16 | 0.3 | 0.5 |
| 3-indoleacetic acid | 1.1 | 0.92, 1.32 | 0.3 | 0.5 | 1.18 | 0.97, 1.43 | 0.093 | 0.2 |
| N-acetylserotonin | 1.05 | 0.96, 1.14 | 0.3 | 0.5 | 0.96 | 0.88, 1.06 | 0.4 | 0.5 |
| 5-methoxytryptophol | 1.09 | 0.87, 1.35 | 0.5 | 0.6 | 1.29 | 1.04, 1.59 | 0.02 | 0.1 |

Cox regression models were stratified by sites and adjusted by age, sex, and GCS. <sup>1</sup> HR = Hazard Ratio, CI = Confidence Interval. <sup>2</sup> Benjamini & Hochberg correction for multiple testing.

**Supplementary Table 2.**
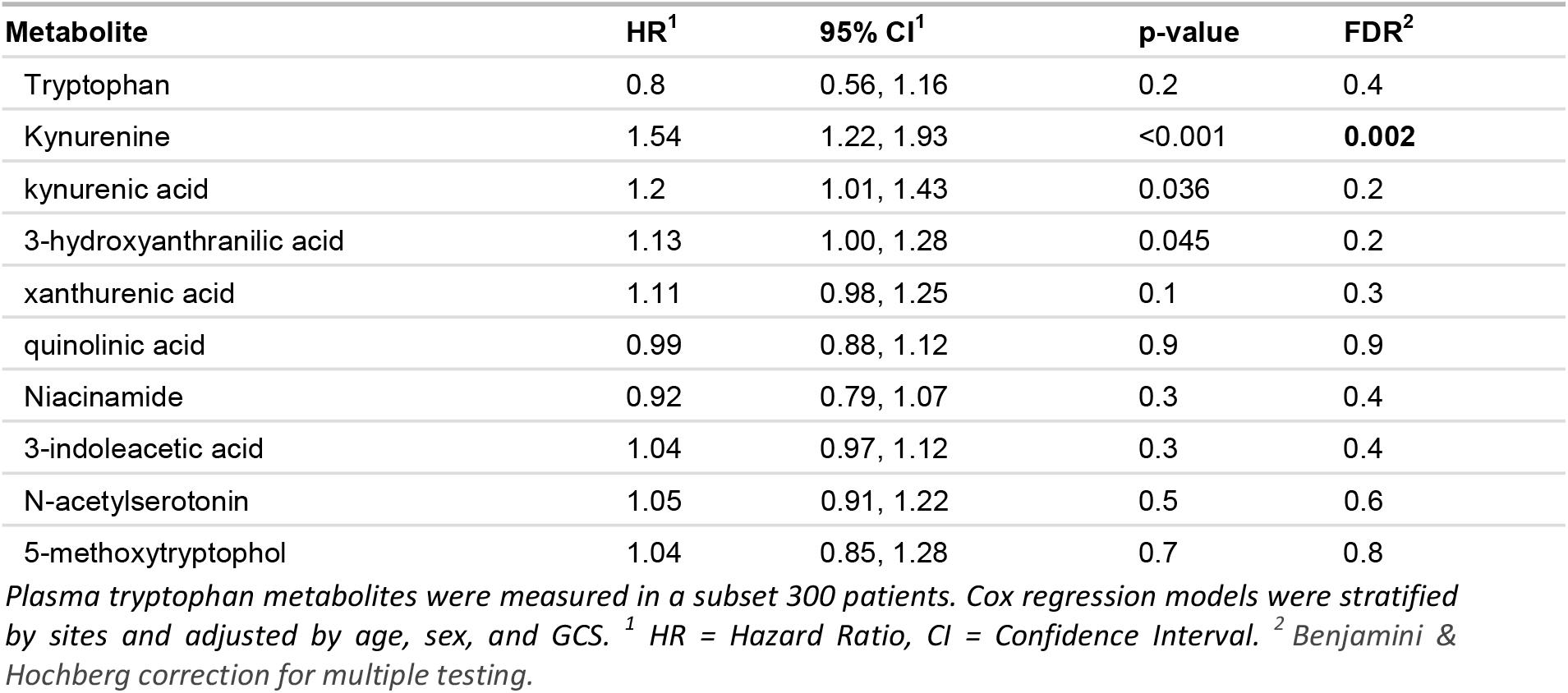
Univariate Cox regression for influence of plasma metabolites on 60-day mortality.

| Metabolite | HR<sup>1</sup> | 95% CI<sup>1</sup> | p-value | FDR<sup>2</sup> |
| --- | --- | --- | --- | --- |
| Tryptophan | 0.8 | 0.56, 1.16 | 0.2 | 0.4 |
| Kynurenine | 1.54 | 1.22, 1.93 | <0.001 | **0.002** |
| kynurenic acid | 1.2 | 1.01, 1.43 | 0.036 | 0.2 |
| 3-hydroxyanthranilic acid | 1.13 | 1.00, 1.28 | 0.045 | 0.2 |
| xanthurenic acid | 1.11 | 0.98, 1.25 | 0.1 | 0.3 |
| quinolinic acid | 0.99 | 0.88, 1.12 | 0.9 | 0.9 |
| Niacinamide | 0.92 | 0.79, 1.07 | 0.3 | 0.4 |
| 3-indoleacetic acid | 1.04 | 0.97, 1.12 | 0.3 | 0.4 |
| N-acetylserotonin | 1.05 | 0.91, 1.22 | 0.5 | 0.6 |
| 5-methoxytryptophol | 1.04 | 0.85, 1.28 | 0.7 | 0.8 |

Plasma tryptophan metabolites were measured in a subset 300 patients. Cox regression models were stratified by sites and adjusted by age, sex, and GCS. <sup>1</sup> HR = Hazard Ratio, CI = Confidence Interval. <sup>2</sup> Benjamini & Hochberg correction for multiple testing.

## Notes

### Competing Interest Statement

The authors have declared no competing interest.

### Author Declarations

Ethical Committee of Hasan Sadikin Hospital, Faculty of Medicine, Universitas Padjadjaran, Bandung, Indonesia Oxford Tropical Research Ethics Committee in the United Kingdom Institutional Review Boards of the Hospital for Tropical Diseases Pham Ngoc Thach Hospital in Vietnam

